# Parvovirus B19 genomic DNA concentrations in wastewater solids are associated with community infections

**DOI:** 10.1101/2024.12.21.24319493

**Authors:** Alessandro Zulli, Rebecca Y. Linfield, Dorothea Duong, Bridgette Shelden, Alexandria B. Boehm

## Abstract

We assessed concentrations of parvovirus B19 DNA from two wastewater treatment plants in a county with a known outbreak in 2024. Wastewater viral concentrations correlated significantly with clinical cases, demonstrating wastewater’s potential for tracking parvovirus B19 infections. Peaks in wastewater concentrations were aligned with the peak in hydrops fetalis diagnoses.

## Introduction

Parvovirus B19 is a single-stranded DNA virus transmitted through respiratory droplets. In 2024 infections increased significantly across the United States, with seroprevalences in children rising from below 3% in 2020-2022 to 24.9% in 2024^1^ It typically causes a mild “slapped cheek” rash in children, but can lead to serious complications in pregnant women, including hydrops fetalis and fetal death.^2–4^ Individuals infected with the virus, particularly those who are immunocompromised, are at risk of developing aplastic anemia.^3,5^

Parvovirus B19 is highly infectious, with an estimated R_0_ of 8, and studies have shown that 20-50% of susceptible people are infected during outbreaks in schools.^6,7^ There is no vaccine available, and limited treatment options, meaning prevention and early detection are crucial.^1,4,8^ However, parvovirus B19 infection is not a notifiable condition to public health authorities in the U.S.^6^ Wastewater based epidemiology has been shown to be an effective, real-time, and low-cost method of generating epidemiological data for a variety of human pathogens, suggesting potential utility for parvovirus B19 .^9,10^

This study aimed to evaluate whether wastewater surveillance can track parvovirus B19 infections by analyzing samples from two wastewater treatment plants during a known outbreak in Montgomery, Texas, and comparing results with clinical information from electronic health records systems.

## The Study

Twenty-four hour composite raw wastewater influent samples were collected three times per week between 18 December 2023 and 30 August 2024 from two wastewater treatment plants serving 65,000 and 70,000 people (WWTP 186 and 187, Figure S1). A total of 220 samples were collected. From each sample, wastewater solids were collected from the sample using settling and centrifugation.

We used a previously validated hydrolysis-probe PCR assay targeting the non-structural protein 1 (NS1) gene of parvovirus B19. Assay specificity and sensitivity were confirmed through in-silico analysis against reference genomes and in-vitro testing against common respiratory viruses and bacteria (appendix, Table S1). DNA was extracted from wastewater solids using the Chemagic Viral DNA/RNA 300 kit followed by column-based PCR inhibitor removal. Viral concentrations of parvovirus B19 (FAM) and SARS-CoV-2 (HEX) were quantified as a duplex assay in six replicate wells using digital droplet reverse transcription polymerase chain reaction (ddPCR, appendix Table S2). We included negative and positive extraction and PCR controls. The detection limit was approximately 1,000 copies per gram of dry weight (cp/g). Primers and thermocycling conditions are provided in the appendix, along with further details of various controls including controls to confirm that storage of samples did not affect nucleic-acid integrity and measurements of pepper mild mottle virus (PMMoV). Measured concentrations in the samples are available through the Stanford Digital Repository (https://doi.org/10.25740/zn011jk5743).

Clinical case and syndromic data was gathered from Epic Cosmos (Epic Cosmos, Epic Systems Corporation, Wisconsin). First, all encounters in the dataset were geographically and temporally filtered to encounters within Montgomery County, Texas between 16 October 2023 and 16 October 2024. The county has a total population of 620,443 as of the 2020 census. From this subset, parvovirus cases and hydrops fetalis diagnoses were identified using grouped International Classification of Diseases (ICD-10) codes.^11^ We identified parvovirus cases using ICD-10 codes B08.3, B34.3, and B97.6, and hydrops fetalis using codes P56.*, O36.*, and Z36.81. Data was aggregated weekly for parvovirus cases and quarterly for hydrops fetalis diagnoses, with values <10 redacted.

Statistical analyses used Kruskal-Wallis tests to compare viral concentrations between plants and chi-square tests for detection frequencies. We assessed associations between wastewater concentrations and clinical cases using Kendall’s tau correlation. Due to multiple comparisons (n=11), we used p=0.005 as the significance threshold.

Parvovirus B19 DNA was detected in 45/111 (40%) of samples from WWTP186 and 62/109 (57%) from WWTP187, with detection frequencies not significantly different (chi-square=5.24, p=0.02). Concentrations were mostly non-detectable before April, peaked in June and returned to non-detectable levels by August. Median (interquartile range, IQR) concentrations were 0 (0-8,071 cp/g) at WWTP186 and 6,121 (0-20,998) copy (cp)/g at WWTP187, showing significant differences between plants (effect size = 6121 cp/g, p=0.001) even when normalized by PMMoV ((effect size = 2.7×10^−5^, p=0.003).

Clinical parvovirus cases in Montgomery County showed similar seasonality, with ≤10 cases until April, peaking in June, and declining by August. Weekly viral concentrations correlated significantly with case counts at both plants (Kendall’s tau=0.56 and 0.52, both p<0.0001). During the quarter of increasing wastewater concentrations, parvovirus cases rose from <10 to 223, coinciding with an increase to 49 hydrops fetalis cases (Figure 1).

**Figure 1.**
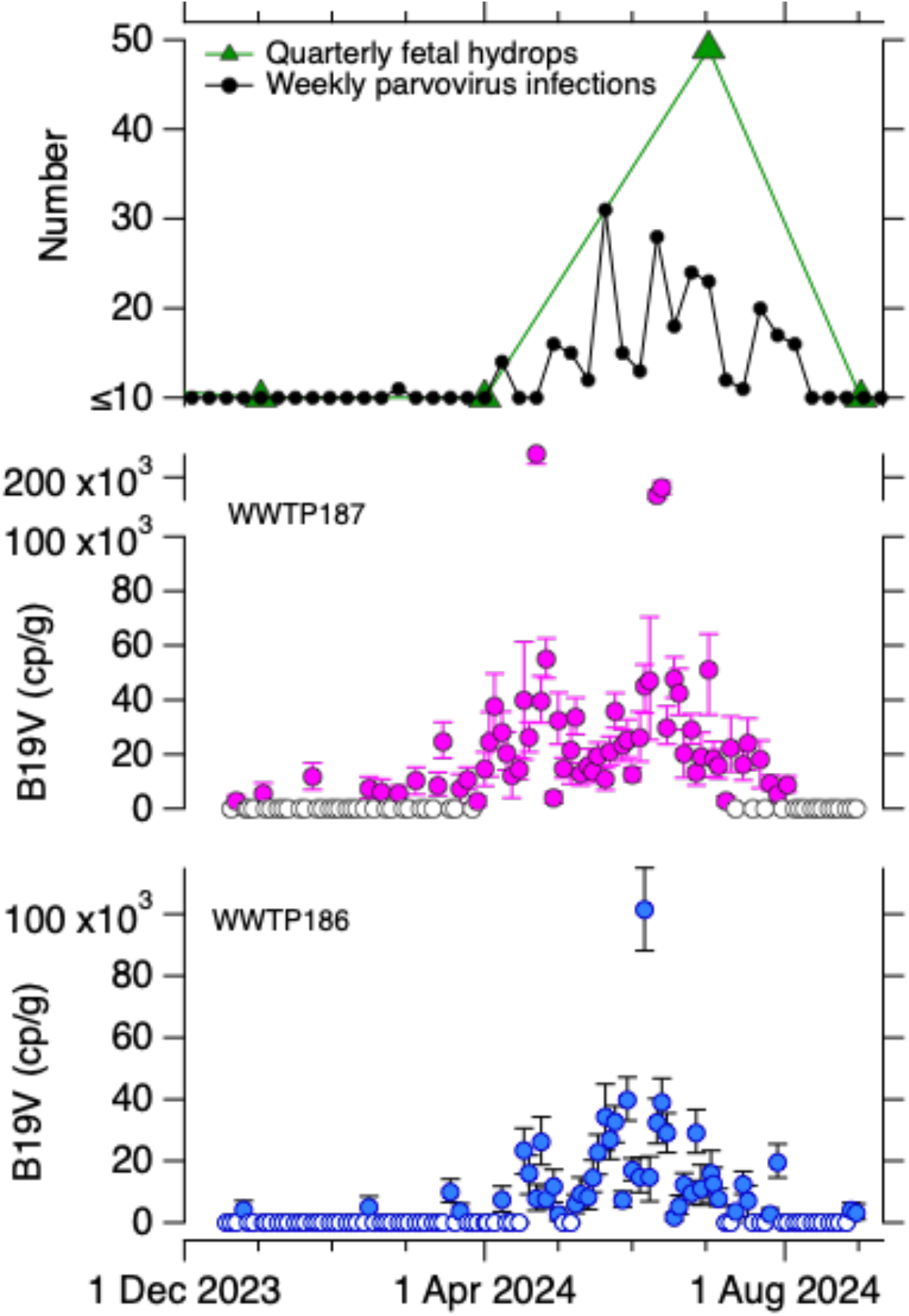
Concentrations of parvovirus B19 (B19V) DNA in wastewater solids in units of copies per gram dry weight at the two indicated WWTP (bottom two panels) and the number of confirmed parvovirus infections and fetal hydrops logged into EPIC for the county where the WWTPs are located (top panel). Open circles in the bottom two panels indicate that B19V was not detected while solid circles indicate it was detected. Error bars represent standard deviations. Note the broken y-axis in the middle panel. For the case and syndromic data, numbers less than or equal to 10 is redacted and reported as ≤ 10. Data on fetal hydrops are provided quarterly; parvovirus infection data are provided weekly. Kendall’s taus between weekly cases and wastewater concentrations were 0.56 and 0.52, respectively (p<0.0001).

This study was reviewed by the Stanford University Committee for the Protection of Human Subjects and determined to be Exempt from oversight.

## Conclusions

This study demonstrates that parvovirus B19 DNA can be quantified in wastewater and directly correlated to clinical indicators of infection. This demonstrates the potential for wastewater to provide sentinel surveillance during key early periods of an outbreak, especially in the absence of mandated public health reporting. Previous studies have shown that parvovirus B19 infects the intestinal mucosa and is present in the sputum of infected patients, indicating it could be shed into wastewater.^12^ Related viruses, such as human bocaviruses, have shown to be shed in stool samples.^13^ Recent studies, such as Li et al., have identified parvovirus B19 (often referred to as erythroparvovirus), in wastewater samples through shotgun sequencing and metagenomic analysis.^14,15^ To our knowledge, ours is the first study to directly quantify parvovirus B19 in wastewater through targeted molecular techniques, and relate the results to clinical case data.

Parvovirus B19 DNA concentrations in wastewater at both treatment plants were significantly correlated with clinical cases of parvovirus B19 in the county. The majority of these diagnosed cases were in children and adolescents under 16 (>90%), likely due to the unique symptomatic presentation in children as a “slapped-cheek” rash and clinical diagnosis without further molecular diagnostics. We then showed that these concentrations corresponded to an increase in maternal care for hydrops fetalis, one of the known complications of parvovirus B19 in pregnant women. This suggests that parvovirus B19 DNA in wastewater is an indicator of not only infections in children, but infections in the adult population, for which clinical data is highly limited. As a result, wastewater represents a rapid and cost-effective method of providing real-time information on parvovirus B19 outbreaks in populations. This is especially important because children are no longer viremic by the time the characteristic rash appears. Early increases in wastewater detection of viremia could then lead to mitigation strategies such as warning or screening pregnant women and those who are immunocompromised.

## Data Availability

Data are available through the Stanford Digital Repository (https://doi.org/10.25740/zn011jk5743).

https://doi.org/10.25740/zn011jk5743

## Acknowledgments

We thank the participating wastewater treatment plants for their samples for the project. Data used in this study came from Epic Cosmos, a dataset created in collaboration with a community of Epic health systems representing more than 284 million patient records from over 1,500 hospitals and 36,000 clinics from all 50 states, D.C., Lebanon, and Saudi Arabia.

## Notes

### Competing Interest Statement

DD and BS are employees of Verily Life Sciences LLC

### Funding Statement

This work was supported by a gift to ABB from the Sergey Brin Family Foundation. RYL was supported by National Institutes of Health (NIH) Institutional National Research Service Award T32 AI007502.

