## Appendix for "Parvovirus B19 genomic DNA concentrations in wastewater solids are associated with community infections"

#### Methods

**Site and sample collection.** Two wastewater treatment plants (WWTPs) serving Montgomery County, Texas were selected for the study. The WWTP186 and WWTP187 serve 65,000 and 70,000 people, respectively (Figure 1). The locations were chosen because Montgomery County recorded a large number of parvovirus infections during the study period based on data from Epic Cosmos (see below). Fifty milliliters of 24-hour composite raw wastewater influent samples were collected using sterile containers by WWTP staff approximately three times per week between 18 December 2023 and 30 August 2024. Samples were sent at 4°C to the laboratory where they were processed immediately. Time between sample collection and receipt at the lab was typically between 1-3 days, during this time, we expect limited degradation of the RNA targets[1]. A total of 220 unique samples were processed. At the lab, the wastewater solids were collected from the influent by settling for 10–15 min, and using a serological pipette to aspirate the settled solids into another tube.

**Assay specificity and sensitivity testing.** We used a previously developed hydrolysis-probe PCR assay for parvovirus B19[2] (hereafter referred to as "B19V") that targets the gene for non-structural protein 1 (NS1). To ensure assay specificity and sensitivity, we tested the assay in silico, and in vitro against viruses, bacteria, and synthetic genomes (Table 1). Nucleic-acids were extracted and purified from intact viruses or bacteria as described below for the wastewater solids samples and then used neat as template in droplet digital the 1-step RT-PCR assay. The assay was run in a single well using the cycling conditions and post processing using a droplet reader as described below for the wastewater samples in singleplex. Synthetic parvovirus B19 genomic DNA (ATCC VR-3281SD) was used as a positive control.

For in silico analysis, the primers and probes were first compared to the reference genome (NC_000883.2) from National Center for Biotechnology Information (NCBI) to ensure 100% alignment. After this, genomes from1 January 2024 to 1 October 2024 were downloaded from NCBI Virus (n=277) and a consensus genome was generated. The assay was then checked against this consensus genome, as well as a subset of individual genomes from the above list. After this, the primers and probes were run through NCBI Blast, excluding Erythroparvovirus primate1 (synonymous with parvovirus B19) and parvovirus B19, to identify potential off-target hits.

**Table S1. Viruses used to test specificity are indicated as "non target testing" and viruses used positive controls are indicated as "target testing". All non-target controls are sold by Zeptomatrix (Buffalo, NY), ATCC (American Type Culture Collection, location), and TWIST (South San Francisco, CA). All the viruses from Zeptometrix are included in the NATtrol™ Respiratory Verification Panel (Catalog #: NATRSP-BIO).**

| Virus | Genomic target | Non-target testing (negatives) | Target testing (positives) |
|---|---|---|---|
| Parvovirus B19 | Non-structural | Parainfluenza 1 (NATRSP-BIO, Zeptometrix) | Parvovirus B19 |

| | | | |
|---|---|---|---|
| (B19V) | protein 1 (NS1) | Parainfluenza 2 (Zeptometrix)<br>Parainfluenza 3 (Zeptometrix)<br>Parainfluenza 4 (Zeptometrix)<br>Influenza A H1N1pdm (Zeptometrix)<br>Influenza AH1 (Zeptometrix)<br>Influenza AH3 (Zeptometrix)<br>Influenza B (Zeptometrix)<br>Adenovirus 1 (Zeptometrix)<br>Adenovirus 3 (Zeptometrix)<br>Adenovirus 31 (Zeptometrix)<br>Rhinovirus Type 1A (Zeptometrix)<br>RSV A (Zeptometrix)<br>RSV B (Zeptometrix)<br>SARS-CoV-2 (Zeptometrix)<br>*M. pneumoniae* (Zeptometrix)<br>*C. pneumoniae* (Zeptometrix)<br>Metapneumovirus 8 (Zeptometrix)<br>Coronavirus HKU-1 (Zeptometrix)<br>Coronavirus 229E (Zeptometrix)<br>Coronavirus NL63 (Zeptometrix)<br>Coronavirus OC43 (Zeptometrix)<br>*B. parapertussis* (Zeptometrix)<br>*B. pertussis* (Zeptometrix)<br>Synthetic Influenza B RNA (Twist #103003)<br>Synthetic Influenza A H1N1 RNA (Twist #103001)<br>Synthetic Influenza A H3N2 RNA (Twist #103002)<br>Genomic SARS-CoV-2 gRNA (ATCC VR-1986D) | DNA (ATCC VR-3281SD) |

**Solids pre-analytical methods.** Samples were further dewatered by centrifugation, and dewatered solids were suspended in DNA/RNA Shield (Zymo Research, Irvine, CA) at a concentration of 0.75 mg (wet weight)/ml. The DNA/RNA shield was spiked with bovine coronavirus (BCoV) vaccine as a RNA recovery control. This concentration of solids in buffer has been shown to alleviate inhibition in downstream RT-PCR [3]. A separate aliquot of dewatered solids was dried in an oven to determine dry weight. RNA was extracted from 6 replicate aliquots of dewatered settled solids suspended in the DNA/RNA Shield, and then it was subsequently processed through an inhibitor removal kit . The pre-analytical methods are also provided on protocols.io[3].

**RNA extraction and purification.** RNA extraction and purification was done using the Chemagic Viral DNA/RNA 300 kit H96 for the Perkin Elmer Chemagic 360 (Perkin Elmer, Waltham, MA). It was followed by PCR inhibitor removal with the Zymo OneStep-96 PCR Inhibitor Removal kit (Zymo Research, Irvine, CA). 300 µl of the suspension entered into the nucleic-acid extraction process and 50 µl of nucleic-acids are retrieved after the inhibitor removal kit.

**Digital droplet RT-PCR analytical methods.** Each replicate RNA extract from each sample (6 per sample) was processed to measure human viral nucleic-acid concentrations using digital

RT-PCR, each in its own well (6 replicate wells per sample). We quantified the number of copies of B19V DNA using the previously established assay (Table 2). The assay was run in duplex using the probe-mixing approach along with an assay for the SARS-CoV-2 N gene; the probe used to detect B19V was labeled using FAM (6-fluorescein amidite), and the probe used for SARS-CoV-2 was labeled with HEX (hexachlorofluorescein). Extraction negative (BCoV spiked buffer, 2 wells) and positive (buffer spiked with positive control cDNA of SARS-CoV-2 target, 1 well) controls, and PCR negative (molecular grade water, 2 wells) and positive controls (cDNA, 1 well) were run on each 96 well plate. Nucleic-acids were stored between 2 and 10 months at -80°C prior to these measurements.

ddRT-PCR was performed on 20 µl samples from a 22 µl reaction volume, prepared using 5.5 µl template, mixed with 5.5 µl of One-Step RT-ddPCR Advanced Kit for Probes (Bio-Rad 1863021), 2.2 µl of 200 U/µl Reverse Transcriptase, 1.1 µl of 300 mM dithiothreitol (DTT) and primers and probes mixtures at a final concentration of 900 nM and 250 nM respectively. Primer and probes for assays were purchased from Integrated DNA Technologies (IDT, San Diego, CA) (Table 2. B19V and SARS-CoV-2 nucleic-acids were measured in reactions with undiluted template.

Droplets were generated using the AutoDG Automated Droplet Generator (Bio-Rad, Hercules, CA). PCR was performed using Mastercycler Pro (Eppendforf, Enfield, CT) with the following cycling conditions: reverse transcription at 50 °C for 60 min, enzyme activation at 95 °C for 5 min, 40 cycles of denaturation at 95 °C for 30 s and annealing and extension at 59 °C for 30 s, enzyme deactivation at 98 °C for 10 min then an indefinite hold at 4 °C. The ramp rate for temperature changes were set to 2 °C/second and the final hold at 4 °C was performed for a minimum of 30 min to allow the droplets to stabilize. Droplets were analyzed using the QX200 (Bio-Rad). A well had to have over 10,000 droplets for inclusion in the analysis. All liquid transfers were performed using the Agilent Bravo (Agilent Technologies, Santa Clara, CA).

Thresholding was done using QuantaSoft Analysis Pro Software (Bio-Rad, version 1.0.596) . In order for a sample to be recorded as positive, it had to have at least 3 positive droplets. Replicate wells were merged for analysis of each sample.

These nucleic-acids were also processed immediately without any storage to measure concentrations of pepper mild mottle virus (PMMoV) RNA, BCoV RNA, and SARS-CoV-2 N gene RNA. PMMoV is highly abundant in wastewater globally[4] and is used as an internal recovery and fecal strength control[5]. BCoV RNA is used as an exogenous viral nucleic acid recovery control. The measurement of SARS-CoV-2 N gene RNA on fresh, unstored samples can be compared to that measured in the stored samples (described above) as an indication of nucleic-acid degradation during storage and freeze thaw. Details of these measurements, and the measurements themselves, are published in Boehm et al.[6]

**Table S2. Primer and hydrolysis probes targeting parvovirus B19[2] (B19V) and SARS-CoV-2 N gene[7]. Each probe contained a fluorescent molecule (FAM for B19V, and HEX for the SARS-CoV-2 N gene), as well as ZEN, a proprietary internal quencher from IDT; and IBFQ, Iowa Black FQ.**

| Target | | Sequence |
|---|---|---|
| Parvovirus B19 | Forward | CCACTATGAAAACTGGGCAATA |
| | Reverse | GCTGCTTTCACTGAGTTCTTCA |
| | Probe | AATGCAGATGCCCTCCACCCAG |
| SARS-CoV-2 N gene | Forward | CATTACGTTTGGTGGACCCT |
| | Reverse | CCTTGCCATGTTGAGTGAGA |
| | Probe | CGCGATCAAAACAACGTCGG |

Concentrations of RNA targets were converted to concentrations per dry weight of solids (copies per gram dry weight (cp/g)) using dimensional analysis. The error is reported as standard deviations and includes the errors associated with the Poisson distribution and the variability among the replicates. Three positive droplets across 6 merged wells corresponds to a concentration ~1000 cp/g for solids, respectively). Measured concentrations in the samples are available through the Stanford Digital Repository (https://doi.org/10.25740/zn011jk5743).

**Parvovirus Case and Symptom Surveillance.** Case and syndromic data used in this study came from Epic Cosmos, a dataset created in collaboration with a community of Epic health systems (Epic Cosmos, Epic Systems Corporation, Wisconsin) representing more than 284 million patients from all 50 states, D.C., Lebanon, and Saudi Arabia. Epic Cosmos reflects US population demographics (ref). First, all encounters in the dataset were geographically and temporally filtered to encounters within Montgomery county, Texas between 16 October 2023 and 16 October 16 2024. From this subset, data for parvovirus cases was selected using International Classification of Disease, Tenth Edition (ICD-10) codes B08.3, B34.3 and B97.6 (Erythema infectiosum; Parvovirus infection, unspecified; and Parvovirus as the cause of diseases classified elsewhere).[8,9] The laboratory testing results for parvovirus were not available through Epic Cosmos; however, prior epidemiological studies have also relied on ICD codes for diagnosis given many children are diagnosed clinically without confirmatory laboratory confirmation. Data was then aggregated at the weekly level and exported as CSV files for analysis. Data are redacted for weeks with fewer than or equal to 10 cases. The date assigned to each week is the last day of the week.

We also used quarterly data, defined as January through March and every 3 months following, for parvovirus cases as defined above, and hydrops fetalis. Hydrops fetalis diagnosis drew on 32 ICD-10 codes encompassing hydrops fetalis due to hemolytic disease, maternal care for hydrops fetalis, and encounter for antenatal screening for hydrops fetalis (ICD-10 P56.*; O36.*; Z36.81) . The data assigned to each quarter represents the last day of the quarter.

**Statistics.** All data series were not normally distributed based on the Wilks Shapiro test of normality ($p<0.05$ for case data, B19V, and B19V normalized by PMMoV). Concentrations of B19V and B19V normalized by PMMoV (B19V/PMMoV) were compared between the two WWTPs using Kruskal Wallis tests. We compared the detection of B19V DNA at the two plants using a chi-square test. We tested the hypothesis that weekly median concentrations of B19V and B19V/PMMoV are associated with parvovirus case data using Kendall's tau. Values in the case data < 10 were replaced with 10. We also tested for associations using weekly averages instead of medians. In total, we carried out 11 statistical tests; to account for multiple comparisons, we conservatively used $p = 0.005$ (0.05/11) as cut off for $\alpha = 0.05$.

### Results

**QA/QC.** The previously designed B19V assay was found to be both specific and sensitive, able to detect their intended targets with no cross reactivity. In silico analysis indicated that the assay did not cross react with non-target sequences, and that the assay was able to detect all parvovirus B19. There was no cross reactivity identified in vitro.

Negative and positive extraction and PCR controls on all plates used for environmental sample testing were negative and positive. Median (interquartile range, IQR) BCoV recoveries were 1.1 (0.75, 1.2) for WWTP186 and 0.89 (0.7, 1.1) for WWTP187. Median (IQR) PMMoV were $1.9\times10^{8}$ ($1.2\times10^{8}$ - $2.7\times10^{8}$) cp/g for WWTP186 and $2.2\times10^{8}$ ($1.4\times10^{8}$ - $3.5\times10^{8}$) cp/g for WWTP187 (Figure 2). BCoV recoveries indicate median recoveries close to 100%, and stable PMMoV between and within WWTPs, respectively. This suggests B19V DNA concentrations can be compared over time and between WWTPs as they have similar high recoveries and consistent fecal strength. Median (IQR) ratio of SARS-CoV-2 N gene concentrations in stored versus fresh samples was 1.5 (1.2,1.8) at both WWTP suggesting minimal effect of storage.

**Additional details related to the EMMI guidelines.** Across all the samples run in this study (n=220), the average (standard deviation) number of partitions (droplets) for the across the 6 replicate wells was 89,483 (27,001) for the reaction for duplex B19V and SARS-CoV-2 N gene assays. The volume of the partitions, as reported by the machine vendor is 0.00085 µL. The mean (standard deviation) of copies per partition for each target was $1.04\times10^{-4}$ ($2.37\times10^{-4}$) and $2.57\times10^{-3}$ ($3.11\times10^{-3}$) for B19V and SARS-CoV-2 N gene, respectively. An example fluorescent plot from the QX200 (2 color reader), as well as a spreadsheet version of the EMMI checklist is included in the Stanford Digital Repository with the deposited data (https://doi.org/10.25740/zn011jk5743).

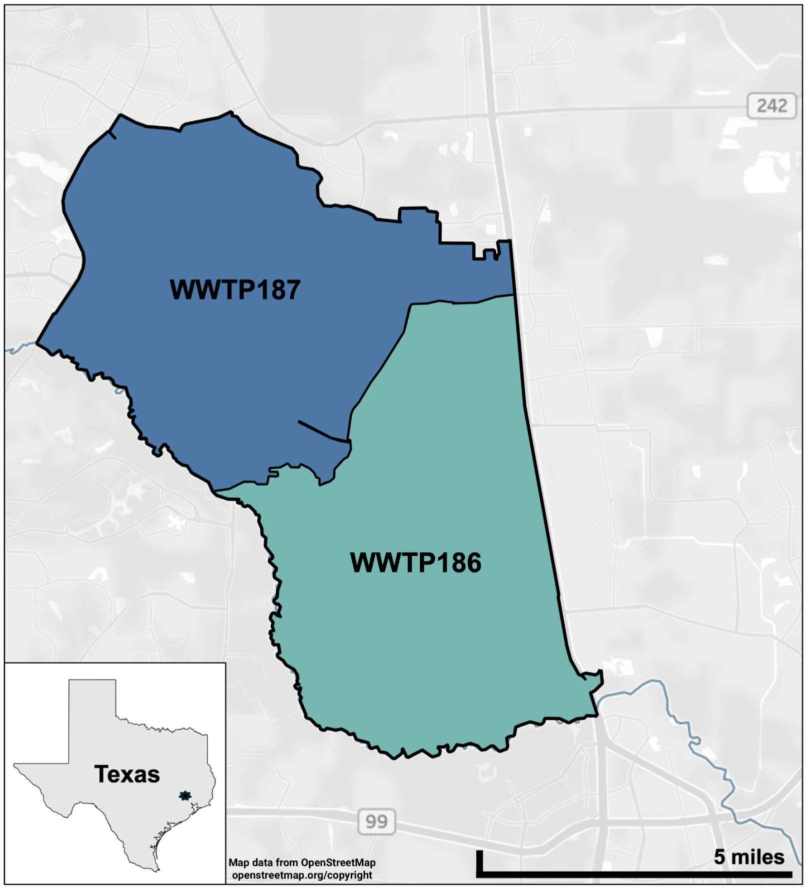


Figure S1. Map of the two sewersheds from which wastewater solids were processed in this study. This figure was generated using Tableau; map layer from OpenStreetMap which is open access(openstreetmap.org/copyright).

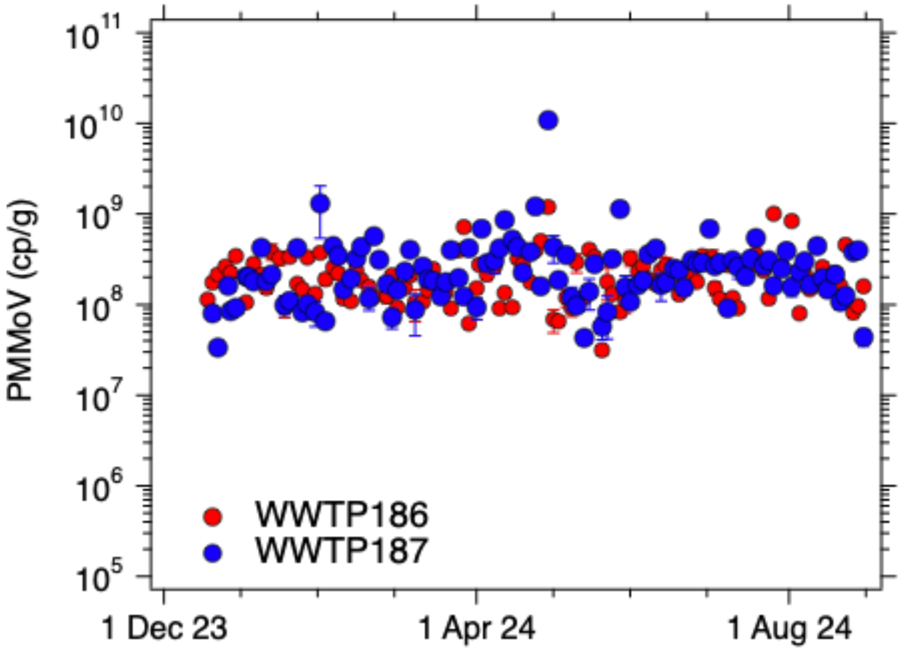


Figure S2. Concentrations of PMMoV measured in the samples, as previously reported[6]. Error bars represent standard deviations and in some cases are too small to be seen.

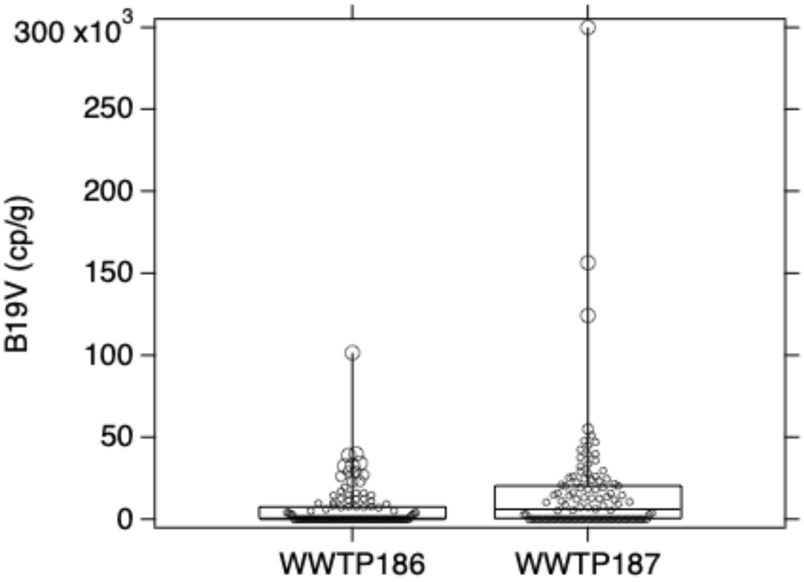


Figure S3. Box and whisker plot of B19V DNA concentrations at the two WWTPs. Lower, middle, and upper lines of boxes represent the 25th, 50th (median), and 75th percentiles. Whiskers extend to minimum and maximum measurements. Data points are shown as circles.

(1) Simpson, A.; Topol, A.; White, B.; Wolfe, M. K.; Wigginton, K.; Boehm, A. B. Effect of Storage Conditions on SARS-CoV-2 RNA Quantification in Wastewater Solids. *PeerJ* **2021**, *9*, e11933. https://doi.org/10.7717/peerj.11933.

(2) Toppinen, M.; Norja, P.; Aaltonen, L.-M.; Wessberg, S.; Hedman, L.; Söderlund-Venermo, M.; Hedman, K. A New Quantitative PCR for Human Parvovirus B19 Genotypes. *J. Virol. Methods* **2015**, *218*, 40–45. https://doi.org/10.1016/j.jviromet.2015.03.006.
(3) Boehm, A. B.; Wolfe, M. K.; Wigginton, K. R.; Bidwell, A.; White, B. J.; Hughes, B.; Duong; Chan-Herur, V.; Bischel, H. N.; Naughton, C. C. Human Viral Nucleic Acids Concentrations in Wastewater Solids from Central and Coastal California USA. *Sci. Data* **2023**, *10*, 396.
(4) Symonds, E. M.; Nguyen, K. H.; Harwood, V. J.; Breitbart, M. Pepper Mild Mottle Virus: A Plant Pathogen with a Greater Purpose in (Waste)Water Treatment Development and Public Health Management. *Water Res.* **2018**, *144*, 1–12. https://doi.org/10.1016/j.watres.2018.06.066.
(5) McClary-Gutierrez, J. S.; Aanderud, Z. T.; Al-faliti, M.; Duvallet, C.; Gonzalez, R.; Guzman, J.; Holm, R. H.; Jahne, M. A.; Kantor, R. S.; Katsivelis, P.; Kuhn, K. G.; Langan, L. M.; Mansfeldt, C.; McLellan, S. L.; Mendoza Grijalva, L. M.; Murnane, K. S.; Naughton, C. C.; Packman, A. I.; Paraskevopoulos, S.; Radniecki, T. S.; Roman, F. A.; Shrestha, A.; Stadler, L. B.; Steele, J. A.; Swalla, B. M.; Vikesland, P.; Wartell, B.; Wilusz, C. J.; Wong, J. C. C.; Boehm, A. B.; Halden, R. U.; Bibby, K.; Delgado Vela, J. Standardizing Data Reporting in the Research Community to Enhance the Utility of Open Data for SARS-CoV-2 Wastewater Surveillance. *Environ. Sci. Water Res. Technol.* **2021**, *7* (9), 1545–1551. https://doi.org/10.1039/D1EW00235J.
(6) Boehm, A. B.; Wolfe, M. K.; Bidwell, A. L.; Zulli, A.; Chan-Herur, V.; White, B. J.; Shelden, B.; Duong, D. Human Pathogen Nucleic Acids in Wastewater Solids from 191 Wastewater Treatment Plants in the United States. *Sci. Data* **2024**, *11* (1), 1141. https://doi.org/10.1038/s41597-024-03969-8.
(7) Huisman, J. S.; Scire, J.; Caduff, L.; Fernandez-Cassi, X.; Ganesanandamoorthy, P.; Kull, A.; Scheidegger, A.; Stachler, E.; Boehm, A. B.; Hughes, B.; Knudson, A.; Topol, A.; Wigginton, K. R.; Wolfe, M. K.; Kohn, T.; Ort, C.; Stadler, T.; Julian, T. R. Wastewater-Based Estimation of the Effective Reproductive Number of SARS-CoV-2. *Environ. Health Perspect.* **2022**, *130* (5), 057011–1.
(8) *ICD-10 Version:2019*. https://icd.who.int/browse10/2019/en (accessed 2024-12-12).
(9) ICD-10 : International Statistical Classification of Diseases and Related Health Problems : Tenth Revision, 2004, Spanish version, 1st edition published by PAHO as Publicación Científica 544.
